# Cerebellar and Prefrontal Structures Associated with Executive Functioning in Pediatric Patients with Congenital Heart Defects

**DOI:** 10.1101/2021.11.09.21266092

**Authors:** Daryaneh Badaly, Sue R. Beers, Rafael Ceschin, Vincent K. Lee, Shahida Sulaiman, Alexandria Zahner, Julia Wallace, Aurélia Berdaa-Sahel, Cheryl Burns, Cecilia W. Lo, Ashok Panigrahy

## Abstract

**Objective:** Children, adolescents, and young adults with congenital heart defects (CHD) often display cognitive and behavioral manifestations of executive dysfunction. We consider the prefrontal and cerebellar brain structures as mechanisms for executive dysfunction among those with CHD.

**Method:** 55 participants with CHD (*M* age = 13.93) and 95 healthy controls (*M* age = 13.13) completed magnetic resonance imaging (MRI) of the brain, from which we extracted volumetric data on prefrontal and cerebellar regions. Participants also completed neuropsychological tests of executive functioning; their parents completed behavioral ratings of their executive functions.

**Results:** Compared to healthy controls, those with CHD had smaller cerebellums and lateral, medial, and orbital prefrontal regions, they performed more poorly on tests of working memory, inhibitory control, and mental flexibility, and their parents rated them as having poorer executive functions across several indices. Across both groups, there were significant correlations for cerebellar and/or prefrontal volumes with cognitive assessments of working memory, mental flexibility, and inhibitory control and with behavioral ratings of working memory, task initiation, and emotional control. Greater prefrontal volumes were associated with better working memory, among those with larger cerebellums (with group differences based on the measure and the prefrontal region). Greater prefrontal volumes were related to better emotional regulation only among participants with CHD with smaller cerebellar volumes, and with poorer inhibition and emotional regulation only among healthy controls with larger cerebellar volumes.

**Conclusion:** The cerebellum modulates the relationships between prefrontal regions and executive functioning differently for pediatric patients with CHD versus health controls.

---

Advances in diagnostic, medical, and surgical techniques have dramatically improved the life expectancy of individuals with congenital heart defects (CHD), particularly among those with complex lesions requiring surgery early in life (Warnes et al., 2001). With improved survival rates, greater attention has turned to the development and quality of life of those with CHD. Historically, clinicians and researchers have focused on anomalies in motor development, as these salient deficits appear early on (Aisenberg, 1982). With medical advances resulting in successive cohorts surviving into adulthood, there was a shift towards evaluating complex cognitive deficits, which are often not evident until school entry or later (Wernovsky, 2006). Empirical work has documented a unique pattern of cognitive sequelae among children and adolescents with CHD (for a review, see Bellinger & Newburger, 2013). Notably, they have an increased risk of deficits in executive functioning, including working memory, task initiation, inhibition of prepotent responses, and shifting between tasks and streams of information.

Beyond cognitive deficits, children and adolescents with CHD often exhibit behavioral expressions of executive dysfunction. For example, their parents often rate their working memory, inhibition, and cognitive flexibility as poorer than typically developing peers (Cassidy, White, DeMaso, Newburger, & Bellinger, 2015; Gerstle, Beebe, Drotar, Cassedy, & Marino, 2016; Hövels-Gürich et al., 2002). Interestingly, work with an array of populations (Anderson, Anderson, Northam, Jacobs, & Mikiewicz, 2002; MacAllister et al., 2012; Mangeot, Armstrong, Colvin, Yeates, & Taylor, 2002; Payne, Hyman, Shores, & North, 2011), including individuals with CHD (Cassidy et al., 2015; Gerstle et al., 2016; Hövels-Gürich et al., 2002)), has consistently documented modest correlations between cognitive and behavioral measures of executive functioning, suggesting that the different assessment methods offer both unique and overlapping information on development. In turn, prior studies have found that cognitive and behavioral measures have both unique and overlapping associations with brain volume and cortical thickness (Faridi et al., 2014; Mahone, Martin, Kates, Hay, & Horská, 2009). Consequently, we examined both cognitive and behavioral measures of higher order skills among children, adolescents, and young adults with CHD and healthy peers.

Whereas the cognitive deficits of children and adolescents with CHD are increasingly well documented, the underlying mechanisms are not. Traditionally, deficits in executive functioning have been attributed to prefrontal dysfunction (Stuss & Benson, 1986). In line with this suggestion, adolescents with CHD have reduced prefrontal volumes (Von Rhein et al., 2014). Diminished white matter connectivity along the precentral sulcus has also been related to higher behavioral ratings of executive dysfunction and symptoms of attention-deficit/hyperactivity disorder (ADHD) among adolescents with CHD (Rollins et al., 2014).

Empirical work has documented that higher order skills are not solely mediated by the prefrontal areas of the brain (Alvarez & Emory, 2006; Stuss & Alexander, 2000). The cerebellum may also contribute to both early motor delays and later higher-order cognitive deficits among those with CHD. There is growing evidence that the cerebellum plays a key role in both cognition and behavior (Rapoport, Reekum, & Mayberg, 2000), including executive functions and behavioral symptoms of ADHD (Bellebaum & Daum, 2007; Gottwald, Mihajlovic, Wilde, & Mehdorn, 2003). The cerebellum has one of the highest regional brain blow flow requirements during the late gestational and early postnatal periods (Chugani, Phelps, & Mazziotta, 1987; Tokumaru, Barkovich, O’Uchi, Matsuo, & Kusano, 1999) and, as such, may be susceptible to growth disturbances among those with CHD, who can be at risk for poor cerebral oxygen and substrate delivery early in life (Donofrio et al., 2003). Indeed, among infants, children, adolescents, and young adults with CHD, neuroimaging studies have found reductions in cerebellar volumes (Semmel, Dotson, Burns, Mahle, & King, 2018; von Rhein et al., 2014, 2015). In turn, cerebellar volumes have been associated with working memory among adolescents with CHD (von Rhein et al., 2014) and inhibition, mental flexibility, and behavioral manifestations of executive functioning among young adults with CHD (Semmel et al. 2018).

Interestingly, theoretical models have suggested that the cerebellum is key for the prefrontal system’s development of higher-order thinking skills. For instance, Koziol, Budding, and Chidekel (2012) argue that executive functions evolved from the need to anticipate and control behavior and that the cerebellum instructs the prefrontal systems on how to plan and problem solve by providing control mechanisms. Recent research similarly suggests that cerebellar development plays a critical role in the organization and development of downstream cortical structures, such as the prefrontal cortex. For example, Limperopoulos and colleagues (2014) found that, among children who were born prematurely (with a mean age of 34 months), cortical growth was inversely related to the degree of early cerebellar injury. Moreover, prefrontal and contralateral cerebellar regions activate in concert while performing executive function tasks (Diamond, 2000). As such, cerebellar anomalies may modulate the role of prefrontal areas on executive functions, either as a function of early development or subsequent coordinated activity.

In the current study, we examined both the unique and interactive associations of prefrontal and cerebellar structures with executive functioning among children, adolescents, and young adults with CHD. We used multiple outcome measures, including paper-and-pencil and computerized tests of cognition and parental ratings of behavior. It was expected that both prefrontal and cerebellar structures would be associated with executive function, and cerebellar volume would moderate associations between prefrontal volumes and executive functioning.

## Methods

As part of a prospective study of brain development among children, adolescents, and young adults with CHD, we recruited 72 participants with varied heart lesions and 99 healthy peers between the ages of 6 and 25 years. Participants were recruited from a single center, using print and digital advertisements, an online registry of healthy volunteers, and referrals from targeted clinics. Study procedures included brain magnetic resonance imaging (MRI), neuropsychological testing, and a review of demographic information and medical records. Study exclusion criteria included comorbid genetic disorders, contraindications for MRI (e.g., a pacemaker), and non-English speakers. For healthy controls, study exclusion criteria also included preterm birth and neurological abnormalities (e.g., brain malformations, strokes, hydrocephalus). In addition to exclusion criteria, 17 participants with CHD and four healthy participants were not included in the final sample, as they did not complete brain imaging, the quality of their imaging was poor, or they did not complete any part of the neuropsychological testing. Our final sample included 55 participants with CHD and 95 healthy peers. Of the final sample, 20 participants with CHD and 54 comparison peers had complete neuropsychological testing; data were not complete for all individuals, as participants elected not to complete parts of the evaluation or were ineligible for portions due to their age. Participants who had reached the age of majority provided informed consent; minors were assented to the project, and their parent or legal guardian provided consent on their behalf. The project was approved by our Institutional Review Board (IRB) and completed in accordance with the ethical principles of the Helsinki Declaration.

Participants underwent brain MRI on a 3 Tesla Skyra scanner (Siemens, Erlangen, Germany), using a 32-channel head coil. 3D sagittally acquired T1-weighted volumetric images were used as input for initial structural segmentation using the standard FreeSurfer pipeline (Fischl, 2012). Automated cortical segmentation was done based on the Desikan Killiany Atlas, which subdivides the brain into 24 discrete cortical regions. For our analyses, we initially aggregated this parcellation into the specific cortical subdivisions of the prefrontal lobe (Figure 1) to examine structural differences between our two groups. In addition, the cerebellar vermis was manually segmented for each participant to obtain finer granularity than is provided by the FreeSurfer templates. The vermis was parcellated into superior, middle, and inferior segments (Figure 2). The total volume for each brain structure was then extracted. In addition, images were reviewed by an experienced pediatric neuroradiologist, and no significant acquired or developmental brain abnormalities were noted in the cerebrum or cerebellum for our sample.

**Figure 1.**
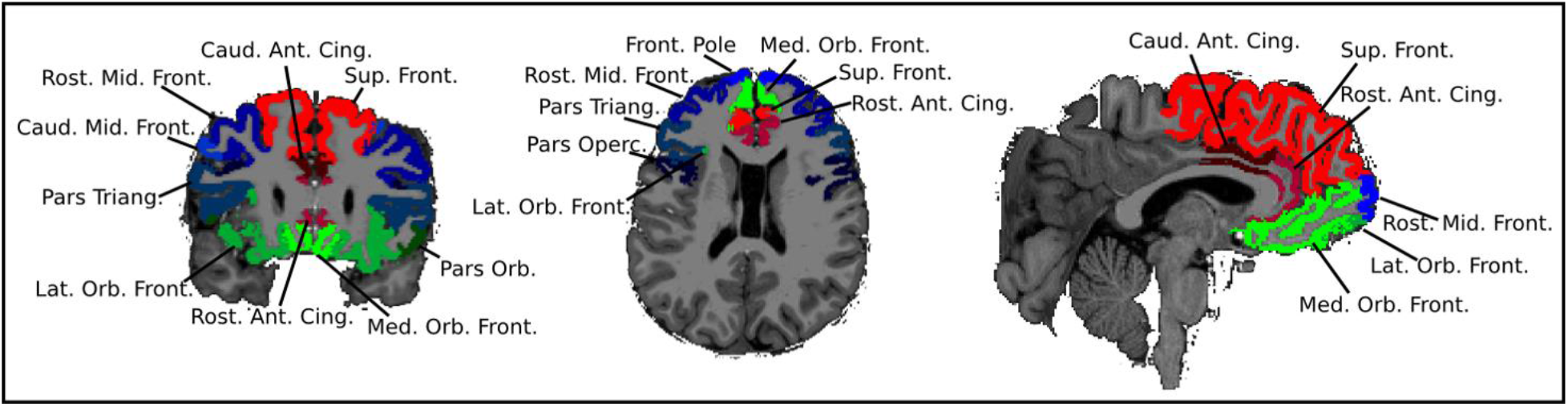
Subdivisions of the Frontal Lobe

**Figure 2.**
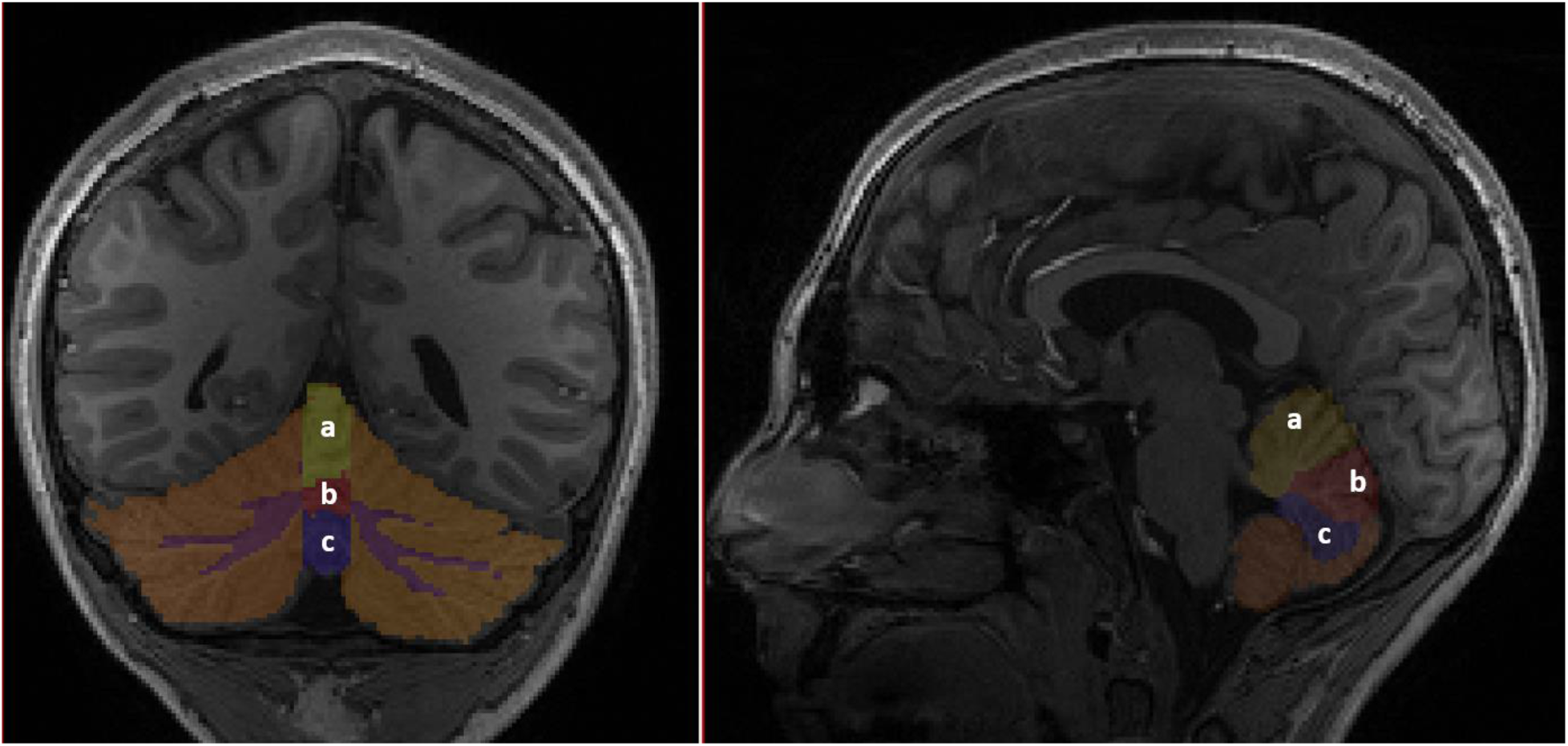
Subdivisions of the Cerebellum *Note*. a. Superior Vermis. b. Middle Vermis. c. Inferior Vermis.

A trained technician supervised by an experienced neuropsychologist administered a battery of neuropsychological tests, which included both clinician- and computer-administered tests, providing multiple assessments of similar constructs. To assess general intellectual functioning, participants of all ages completed the Wechsler Abbreviated Scale of Intelligence, 2^nd^ Edition (WASI-II) (Wechsler, 2011). We considered the four-subtest Full Scale IQ from the WASI-II, a composite of verbal and nonverbal reasoning abilities. To examine executive functioning, we first assessed working memory. Participants between 6 and 16 years old completed the Digit Span and Letter-Number Sequencing subtests of the Wechsler Intelligence Scale for Children Test, 4^th^ Edition (WISC-IV) (Wechsler, 2003); the subtests assess abilities to attend to and manipulate in mind auditory information. Participants ages 7 and up also completed the List Sorting Working Memory Test from the NIH Toolbox (Gershon et al., 2013); the subtest assesses working memory with auditory and visual stimuli. To further investigate executive functioning, participants ages 8 and up completed the Color-Word Interference Test (CWIT), the Trail Making Test (TMT), and the Verbal Fluency Test (VFT) from the Delis-Kaplan Executive Function System Test (D-KEFS) (Delis, Kaplan, & Kramer, 2001); the subtests examine inhibition, sequencing skills, and verbal fluency, respectively, as well as cognitive flexibility. Participants of all ages were also administered the Flanker Inhibitory Control and Attention Test and the Dimensional Change Card Sort Test from the NIH Toolbox, measures of inhibitory control and cognitive flexibility, respectively.

Parents completed behavioral ratings of executive functioning on the Behavioral Rating Inventory of Executive Function Test (BRIEF) (Gioia, Isquith, Retzlaff, & Espy, 2010), which is available for children and adolescents ages 5 to 18. Validity indices on the BRIEF were acceptable for all participants. Higher scores on the BRIEF denote worse functioning.

## Results

Participant demographics are detailed in Table 1. Although participants with CHD had a slightly lower level of intellectual functioning compared to healthy controls, they generally performed within normal limits on tests of intelligence, similar to prior research (Cassidy et al., 2015; Gerstle et al., 2016; Hövels-Gürich et al., 2002). In addition, there was a larger percentage of individuals who identified as White among participants with CHD as compared to healthy controls. Otherwise, the two groups of participants did not differ on demographic variables.

**Table 1.** Demographic Differences Between Youth with CHD and Healthy Controls

|  | Healthy Controls | Youths with CHD | *p* |
| --- | --- | --- | --- |
| Mean Age (SD) | 13.13 (3.75) | 13.93 (4.40) | 0.239 |
| Percent Male | 50.5 | 63.6 | 0.120 |
| Percent White | 62.1 | 87.3 | 0.001 |
| Percent Right Handed | 74.7 | 78.2 | 0.208 |
| Mean WASI-II FSIQ (SD) | 110.65 (12.98) | 104.19 (15.34) | 0.046 |

*Note.* WASI-II FSIQ = Wechsler Abbreviated Scale for Intelligence, 2<sup>nd</sup> Edition Full Scale Intelligent Quotient

We compared the regional prefrontal and cerebellar volumes between patients with CHD and healthy peers using analysis of covariance (ANCOVA) with controls for age and false discovery rate using Benjamini and Hochberg’s (1995) method (Table 2). Participants with CHD had significantly smaller overall cerebellar volume than healthy controls, although there were no significant group differences for the vermis or the regions of the vermis. Prefrontal regions, with the exception of the pars triangularis and the lateral orbitofrontal region, were also significantly smaller among participants with CHD as compared to healthy controls. Next, we examined the bivariate correlations among prefrontal and cerebellar regions. Across the groups of participants, there was structural covariance between total cerebellar volume and each of the prefrontal regions (*r*s *=* 0.29 – 0.53; *p* < 0.05), with the exception of the caudal middle frontal regions among participants with CHD (*r =* 0.12; *p* = 0.39). Among participants with CHD, there was also structural covariance between the middle vermis and the superior frontal region (*r =* 0.28; *p* < 0.05). Among healthy controls, there was structural covariance between each of the cerebellar regions and both the pars opercularis (*r*s *=* 0.23 – 0.43; *p* < 0.05) and the pars orbitalis (*r*s *=* 0.22 – 0.35; *p* < 0.05). Given our interest in the prefrontal regions in relation to cerebellar maturation, we focused our later analyses on the total volumes for the cerebellum, lateral prefrontal region, medial prefrontal region, and orbital prefrontal region rather than subdivisions, based on the initial findings that the overarching regions best captured covariance among the brain structures.

**Table 2.** Cerebellar and Prefrontal Region Structural Differences Between Youth with CHD and Healthy Controls

|  | Healthy Controls |  | Youths with CHD |  |  |
| --- | --- | --- | --- | --- | --- |
| Structure | Mean | SD | Mean | SD | F |
| Cerebellar Regions |  |  |  |  |  |
| Superior Vermis | 6787 | 1305 | 6493 | 1251 | 2.52 |
| Middle Vermis | 4180 | 865 | 3919 | 667 | 4.01 |
| Inferior Vermis | 3643 | 776 | 3775 | 675 | 0.88 |
| Total Vermis | 14610 | 2631 | 14187 | 2281 | 1.36 |
| Total Cerebellum | 108394 | 12552 | 101891 | 11830 | 8.83* |
| Lateral Prefrontal Regions |  |  |  |  |  |
| Pars Opercularis | 8162 | 1192 | 7773 | 1223 | 5.32* |
| Pars Triangularis | 6656 | 1240 | 6794 | 1405 | 0.49 |
| Rostral Middle Frontal | 25229 | 4219 | 23248 | 5031 | 9.80* |
| Caudal Middle Frontal | 7117 | 1338 | 6634 | 1380 | 5.09* |
| Total Lateral Prefrontal | 47163 | 6442 | 44447 | 7597 | 7.94* |
| Medial Prefrontal Regions |  |  |  |  |  |
| Caudal Anteriorcingulate | 6634 | 1380 | 6379 | 1097 | 9.17* |
| Rostral Anteriorcingulate | 5906 | 871 | 5853 | 863 | 6.11* |
| Superior Frontal | 39226 | 4990 | 37167 | 5347 | 7.27* |
| Total Medial Prefrontal | 51458 | 6316 | 48602 | 6444 | 9.08* |
| Orbital Prefrontal Regions |  |  |  |  |  |
| Pars Orbitalis | 2277 | 432 | 2134 | 570 | 4.83* |
| Lateral Orbitofrontal | 1571 | 301 | 1491 | 360 | 2.71 |
| Medial Orbitofrontal | 2660 | 534 | 2504 | 461 | 4.31* |
| Total Orbital Prefrontal | 6508 | 809 | 6130 | 1007 | 8.81* |

\* denotes *F*-tests which were significant at the .05 level with a control for false discovery rate conducted separately for cerebellar regions and frontal regions.
*Note.* Models controlled for age.

We also examined group differences on neuropsychological testing using independent sample t-tests, with a control for false discovery rate (Table 3). Because scores on these tests are standardized by age, we did not adjust for age, as done above. Participants with CHD performed more poorly on a test of working memory from the WISC-IV (Letter-Number Sequencing), a test of inhibitory control from the D-KEFS (CWIT Inhibition), and tests of mental flexibility on the D-KEFS (TMT Number-Letter Switching) and the NIH Toolbox (Dimensional Change Card Sorting). Parental ratings of working memory and other aspects of executive functioning (with the exceptions of inhibition and organization of materials) differed between participants with CHD and comparison peers, such that those with CHD displayed more behavioral dysregulation.

**Table 3.**
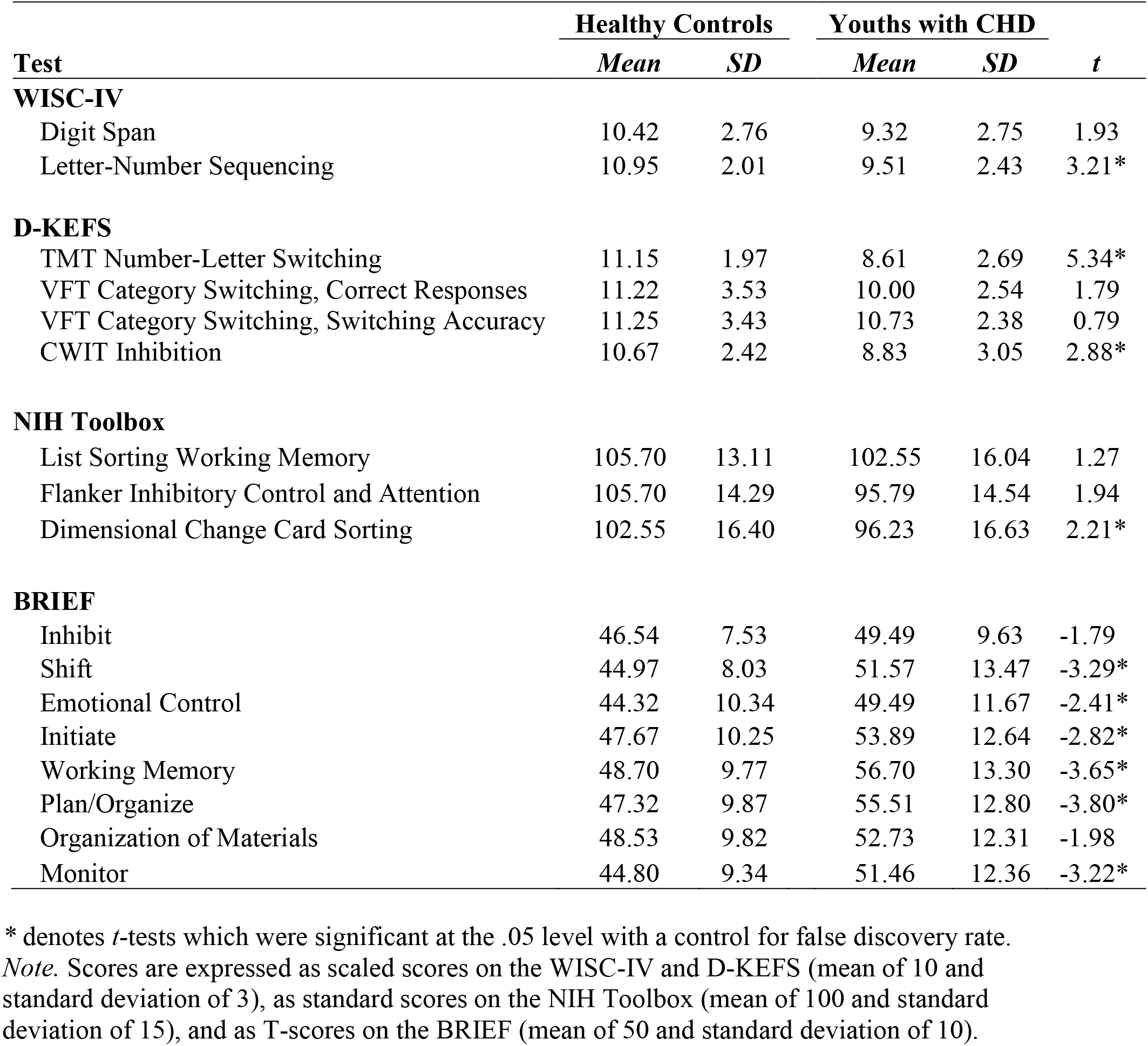
Cognitive and Behavioral Functioning Differences Between Youth with CHD and Healthy Controls

|  | Healthy Controls |  | Youths with CHD |  |  |
| --- | --- | --- | --- | --- | --- |
| Test | Mean | SD | Mean | SD | t |
| **WISC-IV** |  |  |  |  |  |
| Digit Span | 10.42 | 2.76 | 9.32 | 2.75 | 1.93 |
| Letter-Number Sequencing | 10.95 | 2.01 | 9.51 | 2.43 | 3.21* |
| **D-KEFS** |  |  |  |  |  |
| TMT Number-Letter Switching | 11.15 | 1.97 | 8.61 | 2.69 | 5.34* |
| VFT Category Switching, Correct Responses | 11.22 | 3.53 | 10.00 | 2.54 | 1.79 |
| VFT Category Switching, Switching Accuracy | 11.25 | 3.43 | 10.73 | 2.38 | 0.79 |
| CWIT Inhibition | 10.67 | 2.42 | 8.83 | 3.05 | 2.88* |
| **NIH Toolbox** |  |  |  |  |  |
| List Sorting Working Memory | 105.70 | 13.11 | 102.55 | 16.04 | 1.27 |
| Flanker Inhibitory Control and Attention | 105.70 | 14.29 | 95.79 | 14.54 | 1.94 |
| Dimensional Change Card Sorting | 102.55 | 16.40 | 96.23 | 16.63 | 2.21* |
| **BRIEF** |  |  |  |  |  |
| Inhibit | 46.54 | 7.53 | 49.49 | 9.63 | -1.79 |
| Shift | 44.97 | 8.03 | 51.57 | 13.47 | -3.29* |
| Emotional Control | 44.32 | 10.34 | 49.49 | 11.67 | -2.41* |
| Initiate | 47.67 | 10.25 | 53.89 | 12.64 | -2.82* |
| Working Memory | 48.70 | 9.77 | 56.70 | 13.30 | -3.65* |
| Plan/Organize | 47.32 | 9.87 | 55.51 | 12.80 | -3.80* |
| Organization of Materials | 48.53 | 9.82 | 52.73 | 12.31 | -1.98 |
| Monitor | 44.80 | 9.34 | 51.46 | 12.36 | -3.22* |

\* denotes *t*-tests which were significant at the .05 level with a control for false discovery rate.
*Note.* Scores are expressed as scaled scores on the WISC-IV and D-KEFS (mean of 10 and standard deviation of 3), as standard scores on the NIH Toolbox (mean of 100 and standard deviation of 15), and as T-scores on the BRIEF (mean of 50 and standard deviation of 10).

We examined the correlations of the cerebellar volume and the prefrontal volumes (i.e., lateral prefrontal region, medial prefrontal region, and orbital prefrontal region) with neuropsychological functioning (Table 4). Based on Fisher’s *r*-to-*z* transformations, correlations did not statistically significantly differ between participants with CHD and healthy controls; as such, we discuss the pattern of associations across groups. The cerebellum, lateral prefrontal region, and medial prefrontal region were positively associated with working memory on the WISC-IV Letter-Number Sequencing; similarly, the cerebellum and the medial prefrontal region were positively associated with working memory on the NIH Toolbox List Sorting Working Memory. There were positive correlations between the cerebellar, lateral prefrontal, and orbital prefrontal volumes and mental flexibility on the D-KEFS Trail Making Test, and there were positive correlations between each of the prefrontal volumes and mental flexibility on the NIH Toolbox Dimensional Change Card Sorting. Greater cerebellar and prefrontal volumes were associated with greater inhibitory control on both the D-KEFS Color-Word Interference Test and the NIH Toolbox Flanker Inhibitory Control and Attention. On behavioral ratings from participants’ parents, greater cerebellar, lateral prefrontal, and medial prefrontal volumes were related to better task initiation (BRIEF Initiate). In addition, greater cerebellar volume was related to better working memory and emotional control (BRIEF Working Memory, Emotional Control).

**Table 4.**
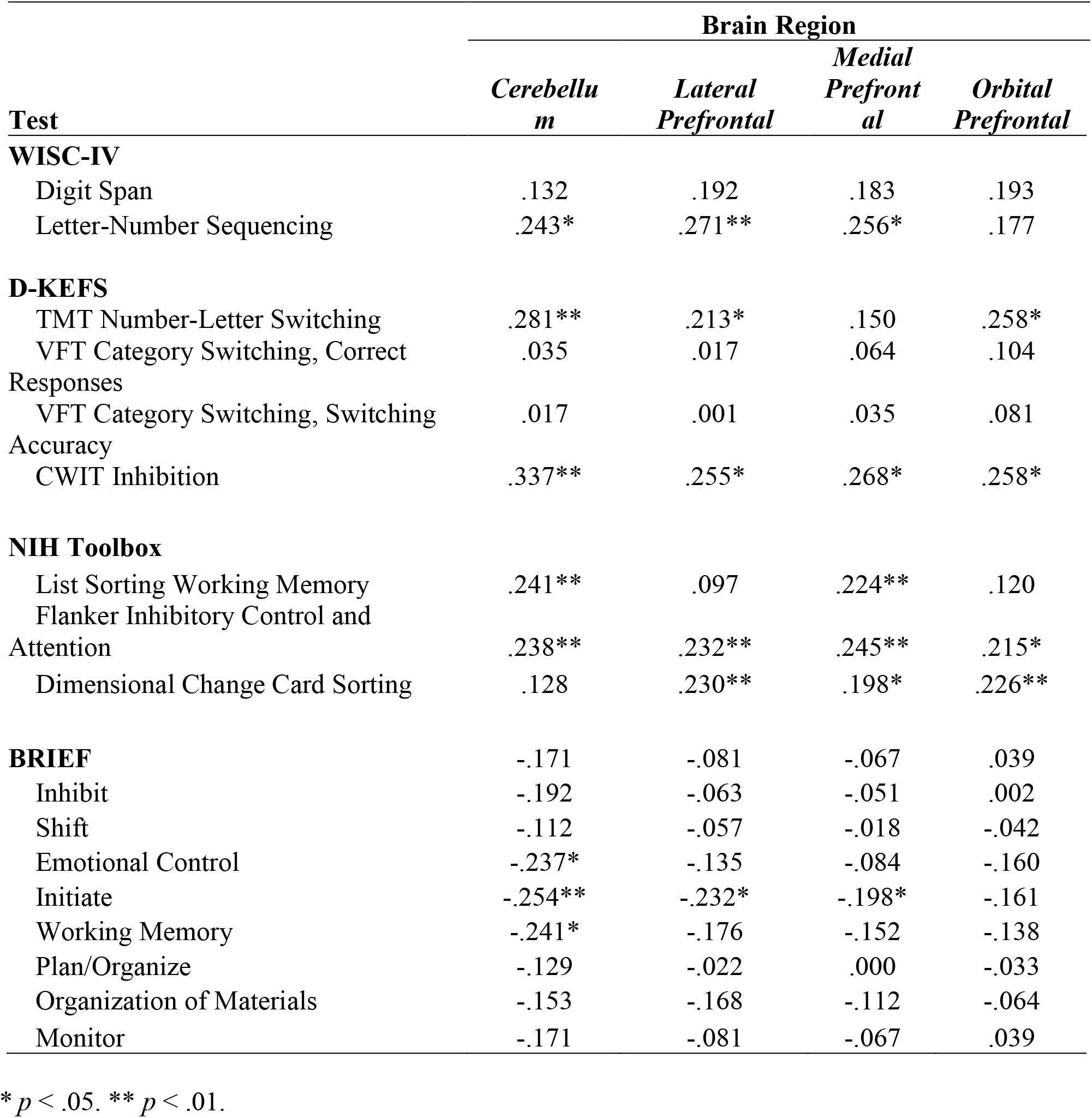
Correlations of Cerebellar and Prefrontal Regions with Cognitive and Behavioral Functioning

Next, we specified models for each cognitive and behavioral outcome examining the interactive effects of cerebellar and prefrontal volumes (Table 5). Main effects for group, cerebellar volume, and prefrontal volume (with separate models specified for each of the three regions) were entered on step 1 of our hierarchical regression models. The interaction for cerebellar volume by prefrontal region was entered on step 2. The three-way interaction for group by cerebellar volume by prefrontal region was then entered on step 3. To decompose any significant interactions with continuous moderators, we followed the recommendations of Aiken and West (1991), testing simple slopes after algebraically fixing the variable to high, median, and low levels (i.e., 1 standard deviation above the mean, the mean, and 1 standard deviation below the mean).

**Table 5.** Interactive Associations of Cerebellar Volume and Prefrontal Volumes with Cognitive and Behavioral Functioning

| Test | Step | Prefrontal Region<br>in Model | $\beta$ | $p$ |
| --- | --- | --- | --- | --- |
| **WISC-IV** |  |  |  |  |
| Digit Span | 2 | Lateral Prefrontal | -3.274 | .028 |
|  | 2 | Orbital Prefrontal | -2.767 | .035 |
|  | 3 | Orbital Prefrontal | -1.263 | .057 |
| Letter-Number Sequencing | 2 | Lateral Prefrontal | -2.478 | .079 |
| **BRIEF** |  |  |  |  |
| Inhibit | 3 | Lateral Prefrontal | -1.028 | .063 |
|  | 3 | Medial Prefrontal | -1.276 | .028 |
|  | 3 | Orbital Prefrontal | -1.544 | .014 |
| Emotional Control | 3 | Lateral Prefrontal | -1.151 | .035 |
|  | 3 | Medial Prefrontal | -0.970 | .093 |
| Initiate | 3 | Lateral Prefrontal | -0.894 | .096 |
| Working Memory | 3 | Lateral Prefrontal | -0.906 | .078 |
|  | 3 | Medial Prefrontal | -0.940 | .085 |
|  | 3 | Orbital Prefrontal | -1.077 | .074 |
| Monitor | 3 | Lateral Prefrontal | -1.040 | .053 |
|  | 3 | Medial Prefrontal | -1.100 | .055 |

*Note.* Only effects significant at the $p < .10$ level are shown. There were no significant effects at the $p < .10$ level on step 2 or 3 for the models with the following outcomes: D-KEFS TMT-Number-Letter Sequencing, VFT- Category Switching, Correct Responses, VFT- Category Switching, Switching Accuracy, and CWIT- Inhibition; NIH Toolbox List Sorting Working Memory, Flanker Inhibitory Control and Attention, and Dimensional Change Card Sorting; BRIEF Shift, Plan/Organize, and Organization of Materials.

Acknowledging that interpretive caution is needed, we discuss interactive regression effects significant at the .01 level. Because detecting significant interaction terms requires vastly greater sample sizes than main effects, considering a less restrictive threshold of significance can help identify meaningful effects with smaller samples. With this caveat in mind, significant interactions emerged for both lateral prefrontal and orbital prefrontal volumes with cerebellar volume for working memory on the WISC-IV Digit Span and for lateral prefrontal volume with cerebellar volume for working memory on the WISC-IV Letter-Number Sequencing. Greater lateral prefrontal volume was associated with better working memory, among participants with larger cerebellar volumes (WISC-IV Digit Span: ß = 0.360; *p* = .022; WISC-IV Letter-Number Sequencing: ß = 0.339; *p* = .023) but not smaller ones (WISC-IV Digit Span: ß = −0.156; *p* = .345; WISC-IV Letter-Number Sequencing: ß = −0.052; *p* = .741). Although there was a similar pattern of findings for orbital prefrontal volume and working memory on the WISC-IV Digit Span at high levels (ß = 0.241; *p* = .087) and low levels (ß = −0.167; *p* = .294) of cerebellar volume, the interaction differed by group. Greater orbital prefrontal volume was related to better working memory, only among healthy controls with larger cerebellar volumes (ß = 0.446; *p* = .029).

There were several significant three-way interactions for group by cerebellar volume by prefrontal region for behavioral ratings of executive functioning, as depicted in Table 5. However, the decomposition of simple slopes only revealed significant results for a subset of interactions, on which we will focus on our discussion. There was a negative relationship between medial prefrontal volume and difficulties with working memory, which decreased in strength from high (ß = −0.514; *p* = .142) to medium (ß = −0.383; *p* = .046) to low levels of cerebellar volume (ß = - 0.251; *p* = .328), only among participants with CHD. Only among healthy controls with larger cerebellar volumes, greater orbital prefrontal volume was associated with greater difficulties with inhibition, (ß = 0.403; *p* = .031), and greater medial prefrontal volume was associated with greater difficulties with emotional regulation (ß = 0.394; *p* = .041). There was a negative association between lateral prefrontal volume and difficulties with emotional regulation, which was only significant among participants with CHD with smaller cerebellar volume (ß = −0.693; *p* = .025).

Lastly, to account for variation in brain volume across the age range, we reran our linear models partialling out age. As there was a similar pattern of findings, we present the unadjusted models. Given the significant difference in intelligence among participants with CHD and healthy controls, we also reran our models with intellectual functioning as a control. Again, similar findings emerged. Because tests of intelligence incorporate aspects of executive functioning (i.e., reasoning and problem solving) and are highly related to measures of executive functions (Diamond, 2013), we presented the models without the control for intellectual functioning (which would statistically remove an aspect of executive functioning from analyses).

## Discussion

Children and adolescents with CHD, particularly those with complex defects, often present with deficits in executive functioning. Traditionally, such deficits have been attributed to prefrontal dysfunction. Increasingly, research has shown that higher order cognitive skills and their behavioral manifestations are not solely mediated by the prefrontal areas of the brain, and the cerebellum may also play an important role. Thus, we examined the unique and interactive associations of cerebellar and prefrontal structures on executive functioning among patients with CHD as compared to healthy peers. Notably, we focused on school age children, adolescents, and young adults, rather than younger children. Because of the dynamic nature of development, assessments conducted with younger children (which may be limited in scope) often have limited predictive validity for later adjustment (McGrath, 2004). The impact of certain deficits may only be apparent among older children, such as late-maturing executive functioning skills (Bellinger et al., 2003) and behavior regulation abilities (Karsdorp, Everaerd, Kindt, & Mulder, 2007).

We first examined the structural differences in the cerebellar and prefrontal regions among patients with CHD and healthy controls. Compared to healthy controls, participants with CHD had smaller cerebellums as well as lateral prefrontal, medial prefrontal, and orbital prefrontal regions. Prior research has similarly found that prefrontal surface area and cerebellar volume is reduced among adolescents and young adults with CHD compared to healthy peers (Semmel et al 2018; von Rhein et al., 2014). We also examined differences in neuropsychological functioning among participants with CHD and healthy controls. Overall, both groups generally performed within normal limits on cognitive measures and behavioral indices of executive functioning. However, those with CHD had poorer executive functioning on certain cognitive measures and worse executive functioning on certain behavioral indices than their healthy peers, similar to prior research (Cassidy et al., 2015; Gerstle et al., 2016; Hövels-Gürich et al., 2002).

We examined the associations of cerebellar and prefrontal structures on cognitive and behavioral outcomes among patients with CHD and healthy controls. The cerebellum is thought to play a role in cognitive and behavioral regulation starting early in life and persisting into childhood and adolescence. Indeed, prior research has found that, among newborns with acyanotic heart lesions, reduced cerebellar volume is related to poorer behavioral state regulation (Owen et al., 2014), and, among adolescents and young adults with CHD, cerebellar volumes are associated with working memory, inhibition, mental flexibility, and behavioral manifestations of executive functioning (Semmel et al. 2018; von Rhein et al., 2014). Similarly, the current study, which included children, adolescents, and young adults with CHD, found that cerebellar volume was associated with cognitive assessments of working memory, inhibitory control, and mental flexibility as well as behavioral ratings of executive functioning (i.e., working memory, task initiation, and emotional control). In line with the extant literature (e.g., Diamond, 2013; Stuss & Benson, 1986), there were also associations for prefrontal regions with cognitive and behavioral assessments of executive functioning. Interestingly, though, studies with patients with CHD have not consistently revealed associations between prefrontal volumes and executive functions (e.g., von Rhein et al., 2014), underscoring the need for a more refined considerations of brain correlates. With this in mind, we examined the interactive associations of cerebellar and prefrontal structures on cognitive and behavioral functioning. Fronto-cerebellar connectivity has long been documented, with the lateral part of the prefrontal cortex connecting to the cerebellum via pontine nuclei and the cerebellum sending projections back to the prefrontal cortex via the dentate nucleus and thalamus (Baillieux, Smet, Paquier, De Deyn, & Mariën, 2008). Given this cortico-cerebellar loop, the cerebellum may play an important role in modulating the relationship between prefrontal regions and executive functioning (Schmahmann, Guell, Stoodley, & Halko, 2019; Clark, Semmel, Aleksonis, Steinberg, & King, 2021). Indeed, across patients with CHD and their healthy peers, our findings suggested that prefrontal volumes are positively associated with working memory, particularly among those with larger cerebellar volumes. In line with our findings, it has been suggested that the cerebellum may be recruited in supporting the prefrontal cortex when tasks require more cognitive resources (e.g., greater working memory) (Clark et al., 2021). Among young individuals, larger cerebellar volumes may translate to greater support for prefrontal networks, which can in turn more efficiently tackle working memory demands.

Prior empirical and theoretical work has suggested that the cerebellum may be related to not only executive functions but also socioemotional processes. Schmahmann proposed the cerebellar cognitive affective syndrome (CCAS) (Schmahmann & Sherman, 1998), which describes the cerebellum’s involvement in not only cognitive processes but also affective and social regulation. Meta-analyses have documented a key role of the cerebellum in emotional processing (Stoodley & Schmahmann, 2008) and social cognition (Van Overwalle, Baetens, Mariën, & Vandekerckhove, 2014), and those with CHD show higher rates of emotional distress (DeMaso et al., 2017) and impairments in social cognition (Calderon & Bellinger, 2015). Research furthermore suggests that dysmaturation and early injury affecting the cerebellum may affect the maturation of neocortical regions and their functional impact on socioemotional processes (Wang, Kloth, & Badura, 2014). In line with this framework, we found that cerebellar volume was associated with emotional regulation, it moderated the relations between prefrontal volumes and emotional regulation, and moderation effects differed across participants with CHD and healthy controls (which likely differed in early cerebellar structure). That being said, future research is needed to better understand the role of the cerebellum not only on emotional functioning but also on social cognition, behavior, and adjustment among patients with CHD.

We described one of the first studies using a computerized assessment of cognitive functioning (namely, the NIH Toolbox) among young individuals with CHD. Prior research has found that measures of cognitive skills, including executive functioning, from the NIH Toolbox have appropriate convergent and divergent validity with traditional paper-and-pencil measures (Zelazo et al., 2013). Given such sound psychometric properties, there has been growing interest in developing and using computerized tools (e.g., CANTAB, CogState, Impact, NIH Toolbox) to screen for cognitive dysfunction and track changes over time among medically complex individuals in clinical and empirical settings (Hardy, Olson, Cox, Kennedy, & Walsh, 2017). While computerized assessments do not provide as detailed information as comprehensive neuropsychological evaluations, they may help identify those who would benefit from such assessment clinically, and they can provide a more temporally and fiscally efficient avenue for researchers. The results of the present investigation, along with emerging work from other research groups (Calderone et al., 2020; Siciliano et al., 2020), suggest that the subtle cognitive deficits seen among patients with CHD may, in part, be detected by a computerized assessment.

## Limitations and Future Directions

The main limitation of our study is its sample size. Although our sample was larger than those found in prior studies examining the role of cerebellar volume on executive functioning among patients with CHD (Semmel et al., 2018; von Rhein et al., 2014), we had limited power to detect effects within complex models. As a result, we elected to use a less stringent index of significance when interpreting the findings of our regression models with interactive effects. We also limited the scope of our analyses. For example, there was limited power to examine the moderating effect of age, although there is a maturation of the prefrontal cortex and cerebellum and a refinement of executive functions throughout childhood and into young adulthood (Arain et al., 2013; Blakemore & Choudhury, 2006). We furthermore did not explore the impact of biological and environmental factors on associations between the cerebellum and cognitive and behavioral functioning. For instance, participants’ type of heart lesion (e.g., single vs. double ventricle; cyanotic vs. acyanotic), their peri-operative complications, and their socioeconomic status might affect both their brain development and functional outcomes. Indeed, differential associations between cerebellar structure and behavioral regulation have been found among newborns with different heart lesions (Owen et al., 2014). It will be important to not only replicate our findings within larger samples but also extend analyses to additional brain regions, confounds, and moderators.

A second limitation of our study is its cross-sectional design. Although a cross-sectional design allows one to draw conclusions about the relations between different brain structures and functional outcomes, it cannot speak to brain development or changes in functioning. It should be noted, though, that few programs have been able to examine longitudinal associations among neuroradiological and neuropsychological findings, given the more recent recognition of the importance of brain development and quality of life among those with CHD. The Boston Circulatory Arrest Study and a handful of others offer notable exceptions (Rollins et al., 2014).

Although our results suggest that the cerebellum may affect the relationship between prefrontal regions and executive functioning among patients with CHD, it is important to note that our project focuses on the structure of the brain rather than connections among different regions. As such, it may be critical for future research to explore how indices of anatomical connectivity (e.g., measured with diffusion tensor imaging) and functional connectivity (e.g., measured with resting state temporal correlations) between the cerebellum and prefrontal regions are associated with cognitive and behavioral outcomes (Skudlarski et al., 2008). Such investigations may be particularly important, as it has been argued that the functional impact of injury and atypical maturation of the cerebellum is more so due to its connections to extracerebellar regions than its intrinsic characteristics (Limperopoulos et al., 2014; Schmahmann & Pandya, 1997).

## Conclusion

In conclusion, we examined the associations of cerebellar and prefrontal structures on executive functioning among patients with CHD as compared to healthy controls. The study included multiple neuropsychological outcome measures, including paper-and-pencil and computerized tests of cognition and parental ratings of associated behaviors. It has previously been highlighted that the cerebellum shares bidirectional connections with the prefrontal cortex, has been implicated executive processes, and is thought to have a refining or modulating role on cognitive functions (Clark et al., 2021). Current results not only echoed that the cerebellum contributes to executive functioning among young individuals with CHD but also provided the first evidence among those with CHD that the cerebellum may modulate the relationship between prefrontal regions and cognitive and behavioral measures of executive functioning.

## Data Availability

All data produced in the present study are available upon reasonable request to the authors.

## Acknowledgements

We would like to thank the children, adolescents, and young adults who participated in our study, their families, Jenna Gaesser, M.D. for her thoughtful comments on a prior version of this manuscript, as well as Nancy Beluk, Michelle Gruss, Christine Johnson, Fern Wasco, and the Department of Radiology at the UPMC Children’s Hospital of Pittsburgh.

This work was supported by the Department of Defense (W81XWH-16-1-0613), the National Heart, Lung and Blood Institute (R01 HL152740-1, R01 HL128818-05), the National Heart, Lung and Blood Institute with National Institute of Aging (R01 HL128818-05 S1), the National Institute of Neurological Disorders and Stroke (K23 063371), the National Library of Medicine (5T15LM007059-27), the Pennsylvania Department of Health, the Mario Lemieux Foundation, and the Twenty Five Club Fund of Magee Women’s Hospital.

The authors declare no conflicts of interest.

